# FoRSHE-X digital health intervention to improve the quality of life during chemotherapy among gynecological cancer survivors in Indonesia: A protocol for a pilot and feasibility study

**DOI:** 10.1101/2024.02.20.24303103

**Authors:** Yati Afiyanti, Dyah Juliastuti, Winnie Kwok Wei So, Ariesta Milanti, Lina Anisa Nasution, Aprilia Dian Prawesti

## Abstract

Most Indonesian gynecological cancer survivors have unmet supportive care needs during chemotherapy, which may lower their quality of life and discontinue the treatment. Digital health intervention can address this issue. This pilot investigation aims to (1) examine the feasibility and acceptability of a Fighting on distRess, Self-efficacy, Health Effects, and seXual issues (FoRSHE-X) intervention and (2) evaluate prospectively the impact of the study implementation on the level of distress, self-efficacy, side effects’ knowledge and management, and sexual quality of life using the RE-AIM (Reach Effectiveness, Adoption, Implementation, and Maintenance) framework. This is a non-randomized pilot and feasibility study. We will recruit women diagnosed with gynecological cancer undergoing chemotherapy to participate in the FoRSHE-X intervention consisting of ten weeks of social media-based education and telecoaching. We will evaluate the primary outcomes of study feasibility and acceptability, and the secondary outcomes of study impacts at three time points with quantitative and qualitative inquiries. We anticipate a minimum of 30 participants to enroll in the study and complete the assessment. We will disseminate results through conferences and peer-reviewed scientific journals. This study will imply whether a definitive trial to evaluate the potential benefits of the FoRSHE-X is viable and how it should proceed. The protocol can aid researchers or nurses in implementing this approach in their study or practice.

## Introduction

Cancer is the second leading cause of mortality in the world, with 10 million cancer-related deaths estimated to occur in 2020 [1]. According to the World Health Organization (WHO), low- and middle-income countries (LMICs) bear the most tremendous burden of cancer [2] More than 70% of the 10 million cancer-related fatalities were in LMICs [2]. In the case of cervical cancer, the disparity is even more prominent, as 90% of new cases and deaths happened in LMICs [1]. Other gynecological cancer types, including ovarian and uterine corpus cancer, also contribute significantly to the burden of cancer in LMICs [1].

Given the immense cancer concerns in many LMICs, reducing premature mortality through promoting well-being has been listed as one of the targets (target 3.4) of the Sustainable Development Goals (SDG) [3]. Therefore, increasing the country’s capacity to provide quality care across the cancer care continuum, including survivorship care, is necessary. Although cancer has been set among the top national catastrophic diseases to prioritize in Indonesia [4], survivorship care is still suboptimal [5].

An individual can be considered a cancer survivor from diagnosis through the balance of his or her life [6]. Earlier studies showed that most Indonesian gynecological cancer survivors had unmet needs, especially regarding information and comprehensive care [7,8]. The survivors reported that they hardly had enough time and opportunity to discuss with the health providers, particularly the nurses, the physical, psychosocial, sexual, and other issues that arose during and after cancer therapy [7,9]. The availability of cancer survivorship educational media in Bahasa Indonesia is low, and most Indonesian cancer survivors also have inadequate health literacy [10,11]. These conditions might hamper the continuity of care and lead to an unanticipated relapse [12]. In addition, unmet needs were also found to be linked to the low quality of life of gynecological cancer survivors in Indonesia [13,14].

In the digital era, many digital devices and platforms, including mobile phones, mobile applications, websites, and wearable devices, are increasingly used in healthcare services [15]. Digital technologies can be harnessed to enhance communication between cancer patients and healthcare providers [16]. Indeed, prior systematic reviews showed promising findings of the digital interventions’ effects in addressing cancer survivors’ unmet health information needs and physical and psychosocial support [17–20]. Digital interventions can serve as an excellent means to assist cancer survivors in managing their disease and treatment side effects, thereby improving their quality of life and the continuity of the therapy [17,20,21]. Digital health interventions were also found to be similarly effective as usual care to improve the quality of life among cancer survivors [21]. Given their broad potential benefits, the WHO has recommended using digital technologies to strengthen the national health system and achieve the vision of health for all [15].

Nursing intervention for gynecological cancer survivors has not yet been well developed to provide comprehensive and culturally sensitive services in Indonesia, incorporating new technology. Gynecological cancer survivors undergoing cancer therapy will have a better prognosis if they can complete their treatment and understand health management during chemotherapy. Throughout therapy, gynecological cancer survivors have multidimensional needs that, if not well anticipated, may result in therapy termination, relapse, or even death [22]. These needs encompass prevention of recurrence, adopting a healthy lifestyle, managing physical symptoms arising from cancer and its therapy, managing psychosocial and sexual issues, and obtaining information related to self-care [23,24].

We are developing a sustainable intervention that embraces digital technologies to overcome the communication barriers between cancer survivors and healthcare providers and to improve self-management for the physical, mental, social, and sexual effects of chemotherapy in the patient’s life. We aim to leverage a digital health intervention to address the insufficiency of survivorship care for Indonesian gynecological cancer survivors by combining social media-based education and telecoaching. Therefore, we propose a digital solution called Fighting on distRess, Self-efficacy, Health Effects, and seXual issues (FoRSHE-X).

We will conduct a study to examine the feasibility and acceptability of the FoRSHE-X intervention using the RE-AIM (Reach, Effectiveness, Adoption, Implementation, and Maintenance) Framework with the specific primary objectives are as follows (1) to examine the eligibility and acceptability of the participants towards the study intervention, (2) to evaluate the feasibility of digital education, telecoaching, and intervention facilitators, (3) to evaluate the process of the implementation, and (4) to evaluate the intervention sustainability or participants’ involvement over time. Additionally, the secondary objective is to evaluate the impact of the study intervention to the level of anxiety, self-efficacy, side effects’ knowledge and management, and sexual quality of life and the participants’ perspectives towards this study intervention.

## Methods

### Design

We will conduct a non-randomized pilot and feasibility study in which all participants will receive the FoRSHE-X intervention. We will adapt the RE-AIM Framework to evaluate the feasibility and pilot study as outlined by O’Brien and colleagues [25] (See Table 1). The RE-AIM dimensions represent multiple tasks of complex multi-components intervention [25]. This feasibility study asks questions: Can a definitive trial to assess the effect of an intervention be done? Should we proceed with it, and how? [26]. A pilot study is a subset of a feasibility study in which the future trial is entirely or partially performed on a smaller scale [26]. Quantitative and qualitative approaches will be used to achieve the study objectives.

**Table 1.** Defining objectives and data collection using RE-AIM Framework.

| RE-AIM Dimension | Objectives | Quantitative Data | Qualitative Data |
| --- | --- | --- | --- |
| **Reach** | To examine the eligibility and acceptability of the participants | Demographic and clinical data<br>Acceptance of the research invitation | Participants' experience and perspective on the acceptability of the recruitment process and involvement in this study.<br>Facilitators' concerns and challenges in recruiting potential participants. |
| **Effectiveness** | To evaluate the impact of the intervention outcomes | Distress Thermometer<br>Chemotherapy side effects<br>Self-efficacy<br>SQoL-F | Participant's experience and perspectives on the effect of FoRSHE-X intervention towards their health |
| **Adoption** | To evaluate the feasibility of digital education, telecoaching, and intervention agents | Complete response to the questionnaire<br>Facilitator's satisfaction | Researcher daily memoing<br>Facilitators' experience toward digital education and telecoaching |
| **Implementation** | Evaluate the process of implementation (usability) | Coaching log book<br>Proof of telecoaching attendance | Participants' desire and challenges to follow the intervention |
| **Maintenance** | Evaluate the intervention sustainability or participants' involvement over time | Participant's diary notes<br>Participant's adherence on telecoaching<br>Follow-up rate<br>Participant retention | Participant's experience and perspectives towards digital education and telecoaching activities |

### Study setting and participants

The study will be initiated at the Dharmais National Cancer Center (DNCC) in Jakarta, Indonesia. It is a comprehensive cancer center providing cancer care, education, research, and data in Indonesia. As a national referral hospital, it serves as the central hub for cancer diagnosis, treatment, and management, serving patients from Jakarta and nationwide. DNCC offers chemotherapy in several units, including the Systemic Therapy Unit, One Day Care Unit, and Inpatient Unit.

We will recruit participants in the DNCC’s One Day Dare Unit and inpatient units. The eligibility criteria will be (1) female aged 18 years or older, (2) being diagnosed with gynecologic cancer (including cervical, ovarian, uterine, vulvar, vaginal, and fallopian tube cancer), (3) receiving chemotherapy at the DNCC, (4) having a smartphone, (5) willing and physically or cognitively able to participate in the study and follow the study procedures, and (6) being able to communicate in Bahasa Indonesia.

On the other hand, patients diagnosed with severe neurological conditions such as unmanaged mental health diagnosis, current metastases to the brain, delirium, and dementia, as well as those with any prior history of receiving chemotherapy or cancer recurrence, will be excluded because these conditions might confound cognitive functioning [27] Patients with severe hearing or visual impairment that would prevent them from communicating and using the educational digital material will also be excluded. Another exclusion criterion is patients who are single, divorced, or widowed with no sexual partner.

Thirty participants will be recruited using a convenience sampling technique to enroll in the study. A sample size of 30 participants is considered sufficient for evaluating the feasibility of a clinical trial and providing preliminary estimates of the effect sizes of the study outcomes [27].

### Outcomes measures

#### Primary outcomes

The primary outcomes will be examined through quantitative and qualitative data on study feasibility and acceptability. The data will cover participant recruitment and retention, the implementation of the intervention, and participant satisfaction.

The research team will maintain a log book to record (1) the number of gynecological cancer patients approached to participate in the study, (2) the number of patients who met the eligibility criteria, (3) the number of patients who sign informed consent, (4) the number of participants who received the intervention and were retained in the study until the final follow-up (week 10), (5) the number of participants who provided a complete response to questionnaires, (6) the number of participants who complete any measures, regardless of their completion rates, and (7) the number of participants who completed the prescribed number of telecoaching sessions.

We will carry out a qualitative evaluation to assess (1) intervention adherence, (2) acceptability of the recruitment processes, intervention design and delivery, and outcome measurements, (3) facilitators and barriers to recruitment and intervention delivery, and (4) participant satisfaction. Qualitative data will be collected through semi-structured interviews with the participants after the study’s completion. An experienced qualitative researcher will conduct the interviews which will be recorded using an audiotape and transcribed by a research assistant.

#### Sociodemographic and clinical variables

We will use a questionnaire to collect participants’ sociodemographic data (i.e., age, education, marital status, employment, ethnicity, and religious affiliation). We will also obtain clinical data at baseline from the participant’s medical record, including gynecological cancer type and stage and cancer treatment details.

#### Secondary outcomes

##### Distress thermometer

Distress is defined by the National Comprehensive Cancer Network (NCCN) as a “multifactorial unpleasant emotional experience of a psychosocial, and or spiritual nature that may interfere with the ability to cope effectively with cancer, its physical symptoms, and its treatment [28]. The NCCN recommended that distress be recognized, monitored, and managed sufficiently and timely [28]. The NCCN Distress Management Panel also developed the Distress Thermometer (DT), which originated from Roth’s study [29], for the initial distress screening.

The DT is a single-item rating scale from 0 (no distress) to 10 (extreme distress) [28]. Participants will be asked to choose the number that best describes the distress level they have experienced over the past week, which could be related or unrelated to cancer. Meta-analyses of studies conducted in Asia and non-English-speaking countries suggested that the optimal cut-off point of DT was four [30,31]. Therefore, participants who score less than four on the DT are noted to experience mild distress [28], whereas a distress level of ≥ for or above indicates a clinically significant distress level [28]. The DT has been adapted and validated by plenty of studies assessing patients with different cancer types in various languages and countries[28], including Indonesia [32]. The Bahasa Indonesia version of DT, which the NCCN has also verified, will be used in this study to measure the distress level of the participants at baseline and follow-up.

##### Knowledge and practice regarding chemotherapy side effects

At baseline and follow-up, the participants will be asked to indicate their knowledge about chemotherapy side effects, how they manage them, and whether they experienced those symptoms in the past week. These outcomes will be measured using the chemotherapy side effect questionnaire, outlined by Almohammadi et al [33] will be adopted for this study. The questionnaire consists of two subsets. The first part assesses participants’ knowledge of 16 common side effects of chemotherapy, such as nausea and vomiting, fatigue, diarrhea and constipation, mouth ulcers and sores, hair loss, and sexual issues [33]. Yes/no responses will be used in this part of the questionnaire. The second part has seven items to examine the practice of side effect management, for example, “I measure my body temperature” and “I avoid eating spicy or fatty foods.” We create four-scale Likert responses (never, seldom, often, and always) in this practice questionnaire. In our prior work, these side effect-related questionnaires have demonstrated their validity and reliability (Cronbach’s alpha of 0.720 and 0.708, respectively).

##### Self-efficacy

Self-efficacy, posited by Bandura as a strong predictor of behavior, is conceptualized as the perceived capability to perform specific actions that are needed to achieve particular goals [34,35]. Numerous studies have shown the role of self-efficacy in improving self-management among patients with chronic diseases, including cancer [36,37].

The self-efficacy for managing chronic disease 6-item scale (SEMCD6) initially developed by the Stanford Patient Education Research Center will be used to measure the participants’ self-efficacy [38]. This questionnaire is brief and easy to administer as it only contains six items with a 1-10 Likert scale ranging from 1 ’not at all confident’ to 10 ’totally confident’ to indicate how confident a patient is in managing the disease symptoms [38] The scale’s total score is the mean score over at least four of the six items [38] A higher score indicates a higher level of self-efficacy [38]. The original version of this SEMCD6 was proven valid and reliable in patients with chronic diseases [38]. Previous studies have shown satisfactory psychometric properties of the several versions of SEMCD6, e.g., Spanish, German, Swedish, and Portuguese [39–41]. This study has assessed the validity and reliability of the Bahasa Indonesia version of SEMCD6. The Cronbach’s alpha value of the SEMCD6 Bahasa Indonesia version is 0.813.

##### Sexual quality of life (SQoL-F)

Sexual quality of life is the primary outcome of our future RCT and will be measured at baseline and follow-up using the Sexual Quality of Life-Female (SQOL-F). SQOL-F was initially designed by Symonds et al. using data sets from women’s health surveys in the United Kingdom and the United States to examine how female sexual dysfunction impacts women’s sexual quality of life [42]. This tool comprises 18 questions that capture sexual self-esteem, emotional well-being, and relationship issues in women [42]. Each item is rated on a six-point Likert scale (“completely agree” to “completely disagree”). Questions numbers 1, 5, 9, 13, and 18 have reversed scoring. Therefore, the total score ranges between 18 and 108 [42]. A higher score indicates a higher level of sexual quality of life [42].

SQOL-F has been translated into some languages, for example, Iranian [43] Portuguese [44], Turkish [45] and Indonesia [46]. In the prior work of this study, the Bahasa Indonesia version of SQOL-F is valid and reliable (Cronbach’s Alpha = 0.751) among Indonesian gynecological cancer survivors.

### FoRSHE-X intervention

The main components and timeframe of the intervention are summarized in Table 2. The FoRSHE-X intervention consists of social media-based education and telecoaching. It will be implemented in two phases: Phase I will last six weeks, and Phase II will last for four weeks.

**Table 2.** Activities and timeframe of the FoRSHE-X digital intervention.

| Phase | Week | Activity | Indicator |
| --- | --- | --- | --- |
| Working Phase I | I | Digital education,<br>Baseline data collection (T0) | Knowledge of chemotherapy side effects (physical, psychological, sexual) |
|  | II | Telecoaching session 1 | Distress management |
|  | III | Telecoaching session 2 | Physical side effects management |
|  | IV | Telecoaching session 3 | Quality of sexual relationship |
|  | V | Telecoaching session 4 | Self-efficacy |
|  | VI | Evaluation (T1) |  |
| Working Phase II | VII | Telecoaching session 5 (*on demand*) |  |
|  | VIII | Telecoaching session 6 (*on demand*) | Side effects management |
|  | IX | Telecoaching session 7 (*on demand*) |  |
|  | X | Evaluation (T2) |  |

#### Social media-based education

We developed digital educational media, video and infographics, based on clinical guidelines [47] to equip patients with valuable insights and strategies for effectively managing the physical, psychological, and sexual issues that may arise during their treatment journey. The educational contents consist of management of distress, pain, fatigue, anemia, risk of falling, hair fall, skin problems, sleep problems, difficulty breathing, bleeding, dehydration, constipation, diarrhea, loss of taste and smell, nausea and vomiting, weight loss, edema, sexual issues, and infertility. Our educational materials have been crafted to be visually engaging, featuring eye-catching graphics paired with clear and concise descriptions in Bahasa Indonesia to ensure accessibility and understanding for all patients. Recognizing the prevalence and accessibility of social media, we have chosen to leverage the power of Instagram, one of Indonesia’s most widely used social media platforms, as the vehicle through which we provide this patient education.

#### Telecoaching

Telecoaching, a purposeful and patient-centered intervention carried out by certified nurse coaches using WhatsApp video calls, is organized around a ten-step procedure designed to empower and support the participants in applying the knowledge they have obtained from our social media-based education to manage the arising problems related to cancer and chemotherapy. The ten steps are (1) building trust and setting the agenda, (2) finding the inner drive, (3) addressing hidden issues, (4) clarifying orientation, (5) defining goals, (6) identifying options, (7) managing barriers, (8) getting support, (9) taking action, and (10) getting feedback.

The first phase lays a solid foundation for the coaching partnership by establishing trust and planning the agenda. The participants’ inner motivation must then be tapped in step two to ignite their desire for proactive self-management. The subsequent step will address concerns that may be hidden and obstruct growth. Step four, clarifying orientation, is crucial in coordinating the patient’s focus with their health objectives. The joint definition of specific objectives and selecting workable solutions to meet those objectives make up steps five and six. In steps seven and eight, the participants will be coached to develop strategies for overcoming obstacles and exploring avenues of external support. Step nine guides patients to take decisive actions after establishing goals and plans. Lastly, receiving feedback in step 10, which brings the coaching process full circle, guarantees that it is dynamic, flexible, and highly responsive to the changing needs and achievements of the individual.

### Data Analyses

Descriptive statistics of participants’ clinical and demographic characteristics and all study outcomes will be reported. The recruitment and retention rates will be computed to determine the feasibility of a full RCT. A future complete RCT will be deemed feasible if the recruitment rate is at least 50% and the retention rate is at least 50%. Response rates to questionnaires, follow-up rates, and intervention adherence rates will also be calculated to provide further evidence for the feasibility of a definitive RCT. For the secondary objective, we will compare the longitudinal quantitative outcomes at three-time points (before, at the middle, and after the interventions) using the paired t-test (for continuous variables) and McNemar’s test (for categorical variables) to evaluate changes at follow-up.

Qualitative data will be analyzed using content analysis by the research team’s qualitative researchers. The team will develop a codebook based on the study objective and framework. The predetermined codes include acceptability, implementation, facilitators, barriers, and satisfaction. A qualitative researcher will code the transcripts once the final codes have been defined and agreed upon. Emerging codes can be added to the codebook during the analysis. After the analysis is finished, a detailed description of the findings will be reported. The qualitative findings will inform the acceptability of the study and the strategies or best practices of the study protocol.

### Governance

The core team for this research consists of nurses with expertise in cancer care. The principal investigator is a professor of nursing who will oversee the study and deliver the telecoaching intervention. The research team members include nurses working in academia and at the DNCC. The nurses at DNCC will help provide a list of patients who may be eligible. In addition, a medical oncology consultant will also be involved in the study implementation process.

### Research Ethics

Before the research activities commence, the participants will be provided with an explanation of the activities to be conducted and their rights, as detailed in the information sheet. The participants will be able to ask questions about any unclear aspects and have the right to participate in this research voluntarily. Subsequently, if a participant agrees to be involved in this study, she will sign the Informed Consent Form.

### Compensation

Participants will receive an internet data plan worth IDR 100,000 per month for the 2.5-month duration of the research activities. Additionally, a compensation of IDR 100,000 will be provided to each participant at the end of the FoRSHE-X digital intervention to offset the time spent participating in this study. Furthermore, the participants who will also be interviewed after the provision of digital education will receive IDR 100,000 each as compensation for transportation and time used during the interview process.

## Discussion

This study will examine whether a definitive trial to evaluate the potential benefits of the FoRSHE-X intervention that combines digital education and nurse-led telecoaching is viable and how it should proceed. Feasibility comprises the practicability of implementing the trial, including participant recruitment and retention, intervention delivery, and data collection. This study will also seek to determine appropriate and acceptable outcome measures to be included in the full RCT. As such, the intervention will be given to everyone who participated in this feasibility and pilot study. Nevertheless, by using this approach, we cannot test the randomization strategy for the subsequent RCT.

Exploring the participants’ perspectives and experiences will also provide valuable insights into the acceptability of the study. We will assess how well the FoRSHE-X intervention was received and whether participants found it suitable and beneficial. Potential challenges, uncertainties, and strategies to improve them will also be explored by the participants, including those who drop out of the study. The qualitative part of the study can also help identify other potential impacts of the intervention and its interaction with the context in which it is implemented.

Study participants being recruited only from one hospital may limit the generalizability of the results. In this situation, the research findings will likely predominantly reflect the characteristics and experiences of the individuals involved in the study. As a result, the findings might not be readily applicable to a population that is more diverse or geographically separated, such as people who reside in rural areas with little access to online resources. Rural communities often face particular difficulties in gaining access to healthcare and technology, which may substantially impact how they respond to digital interventions like the one under study.

Consequently, while the results may be insightful for the hospital’s patient population, they should be considered relevant to a larger, more diverse population, especially those with limited internet access and lower educational levels.

The primary outcome of the FoRSHE-X intervention will be the sexual quality of life.

Using telecoaching to improve the sexual quality of life for participants and their spouses can be a promising approach, although it may come with several possible difficulties. Firstly, discussions about private matters may give rise to privacy issues, limiting participants’ openness and willingness to engage fully in telecoaching sessions. Secondly, participants in coaching programs often encounter various challenges in implementing the coaching recommendations effectively. One potential major challenge is the resistance to change, as people tend to be more comfortable with their existing mindset and habits. Skills and patience may be needed to overcome such resistance. Thirdly, environmental or resource constraints can be another challenge for this study. Particularly for those living in suburban or rural areas, technological obstacles like poor internet connectivity may hinder effective and smooth communication during sessions. We will explore the participants’ perspectives to address the potential challenges of the trial.

The FoRSHE-X digital intervention is expected to enhance the self-efficacy of gynecologic cancer survivors in preventing and addressing obstacles that may hinder them from continuing therapy and preparing for the post-treatment period. This intervention can connect patients with healthcare teams without space and time constraints. The utilization of digital technology is believed to expand the reach of healthcare services and enhance patient participation in their own sustainable self-care. The FoRSHE-X digital intervention has the potential to meet the supportive care needs of gynecologic cancer survivors undergoing chemotherapy.

## Data Availability

N/A

## Acknowledgments

The authors would like to thank the gynaecological cancer participants who will involve in the pilot of this study and Dharmais Cancer Hospital clinicians for their invaluable support and guide for the development and the implementation of this research project.

## Contributorship

YA and DJ researched literature and conceived the study. LAN and ADP were involved in protocol development, gaining ethical approval, patient recruitment, and data analysis for instrument validity testing. AM and DJ wrote the first draft of the manuscript. YA and WKWS reviewed the manuscript. All authors edited and approved the final version of the manuscript

## Ethical approval

The ethics committees of Nursing Faculty, Universitas Indonesia (REC number: KET- 136/UN2.F12.D1.2.1/PPM.00.02/2023) and Dharmais Cancer Hospital (REC number: 259/KEPK/VIII/2023) approved this study.

